# Resuming assisted reproduction services during COVID-19 Pandemic: An Indian experience

**DOI:** 10.1101/2020.09.20.20198143

**Authors:** Padma Rekha Jirge, Sadhana Patwardhan, Dilip Paranjape, Shishir Narendra Jirge, Deepali Atul Bhomkar, Shruti Mahesh Chougule, Anita Salpekar, Deepak Modi

**Affiliations:** Sushrut Assisted Conception Clinic & Shreyas Hospital, 2013 E, 6^th^ Lane, Rajarampuri, Kolhapur – 416 008, Maharashtra, India; Nagpur Test Tube Baby Centre, Nagpur, Maharashtra, Inida; Sushrut Assisted Conception Clinic & Shreyas Hospital, Kolhapur, Maharashtra, India; Molecular and Cellular Biology Laboratory, ICMR-National Institute for Research in Reproductive Health, JM Street, Parel, Mumbai 400 012, India

**Keywords:** COVID-19 pandemic, assisted reproduction, resumption of in-vitro fertilization (IVF), key performance indicators (KPIs), SARS, coronaviruses

## Abstract

**Purpose:** The pandemic of COVID-19 has affected many countries; and medical services including assisted reproductive treatment (ART) have been hampered. The purpose of the study was to assess the preparedness of ART clinics and staff to resume services; patients’ reasons to initiate treatment and key performance indicators (KPIs) of ART labs during pandemic.

**Methods:** Semi-descriptive report of two private in-vitro fertilization (IVF) clinics in Maharashtra, India, when COVID-19 testing for asymptomatic people was unavailable. Time required for replenishing laboratory supplies, and staff preparedness to function under ‘new norms’ of pandemic was documented. Infection mitigation measures at workplace and triaging strategy were evaluated. KPIs of the clinics were evaluated.

**Results:** 30% of the patients consulted through telemedicine accepted or were eligible to initiate treatment upon clinic resumption. Lack of safe transport and financial constraints prevented majority from undergoing IVF, and only 9% delayed treatment due to fear of pandemic. With adequate training, staff compliance to meet new demands could be achieved within a week, but supply of consumables was a major constraint. 52 cycles of IVF were performed including fresh cycles and frozen embryo transfers with satisfactory KPIs even during pandemic. Conscious sedation and analgesia during oocyte retrieval was associated with reduced procedure time and no intervention for airway maintenance, compared to general anaesthesia. Self reported pain scores by patients ranged from nil to mild on a graphic rating scale.

**Conclusion:** This study provides a practical insight and will aid in forming guidelines for resumption of IVF services during pandemic.

## Introduction

COVID-19 pandemic caused by SARS-CoV-2 virus has strained and challenged the healthcare systems of all the affected countries in an unprecedented manner [1-4]. As a mitigation strategy many countries were under lockdown to minimize human-to-human transmission, and prioritize services of healthcare professionals and medical equipment to the care of seriously sick people [4]. All the non-essential medical services were put on hold leading to suspension of medically assisted reproduction / assisted reproductive treatments (MAR/ART) in majority of the clinics across the globe. Except for ongoing cycles or fertility preservation prior to gonadotoxic therapy, the scientific bodies in assisted reproduction advised suspension of initiation of new treatment cycles, including ovulation induction, intrauterine insemination (IUI), in-vitro fertilization (IVF), intracytoplasmic sperm injection (ICSI), embryo transfers, and non-urgent gamete cryopreservation in March 2020 [2,3,5,6]. They also recommended preferential utilization of telemedicine over ‘in-person’ interactions and suspension of non-urgent diagnostic and elective surgeries [2,3,5,6].

Subsequently, as we understood the population dynamics of the spread of virus and successful mitigation strategies, the reproductive medicine societies advocated gradual and judicious resumption of reproductive care services [7-11]. A general framework for restarting ART activities was released based on the principle that clinic staff is triage-negative and only triage negative patients are offered treatments [7-11]. For the triage positive patients, further decisions are based on results of testing for SARS-CoV-2 [9,11]. Many clinics across the globe adopted this framework and resumed ART services.

India went into national lockdown from 25^th^ March 2020 till 31^st^ May 2020. Movement of people and all forms of transport including goods across states were completely prohibited during the initial weeks. While essential medical services were operational, non-essential services were halted. Most IVF clinics spontaneously stopped services at least in the first six weeks. As time dependent relaxations happened based on the scenario across individual states or territories, infertility being a time sensitive disease, resumption of fertility services became a necessity even in India [11,12]. As per the national guidelines, the SARS-CoV-2 testing by RT-PCR was to be offered to symptomatic patients and those requiring emergency medical services [13,14]. There were no specific guidelines regarding availability of the testing services for ART clinics and patients, and they were not easily available in all parts of the county. This raised the need for a clinic centric protocol to reinitiate treatment, taking cognizance of local scenario and regular internal auditing, in addition to observing international recommendations [15,16].

Herein we describe our experience of resuming IVF services during the span of the pandemic in a hot spot zone of India. We aim to address three important primary outcome measures 1) preparedness of clinics to resume functionality 2) characteristics of patients making an informed decision to initiate treatment and 3) key performance indicators (KPIs) of laboratories following resumption of IVF work. Secondary outcome measures were the efficacy and acceptance of conscious sedation and analgesia (CSA) for oocyte pick up (OPU) in comparison to general anaesthesia (GA).

## Materials and Methods

The data is from two private tertiary fertility clinics – Sushrut Assisted Conception Clinic, Kolhapur (Clinic 1) and Nagpur Test Tube Baby Centre, Nagpur (Clinic 2), located 900 km apart, in the state of Maharashtra, India. The study period is from 14/4/20 to 22/7/20. Informed written consent was obtained from couples for planned treatment and for IVF during COVID-19 pandemic. Consent to follow the prescribed code of conduct was obtained from all the team members and patients (Supplementary data).

### Functional preparedness of the clinics

Both clinics independently maintained basic functionality of IVF laboratories during national lockdown. This meant maintaining an uninterrupted supply of laboratory gases, liquid nitrogen (LN_2_), daily logging of quality control measures and maintaining the stock of all consumables above predefined clinic-specific minimum quantity. While suppliers prioritized provision of LN_2_ to IVF clinics during the lockdown period, a special permission from local authorities was needed for transport. Directives provided by Indian Society for Assisted Reproduction (ISAR) facilitated the above. Distributors of laboratory, operating theatre and ultrasound equipment were contacted and any specific advice for protection of the equipment implemented. Time taken to achieve each of these goals was documented. At the beginning of the lockdown, telephonic contact with all patients who were in different stages of preparation for an ART cycle was established. They were counseled about compliance with lockdown rules, and importance of ‘new norms’ (social distancing, wearing face masks in public places and hand sanitization). Further, they were encouraged to follow healthy lifestyle with a combination of exercise and diet and not to visit hospital without prior arrangement.

Simultaneously, steps were initiated towards COVID-19 specific effective functioning of the clinic personnel during the pandemic; and regular ‘mock-drills’ were commenced. These involved adherence to ‘new norms’, undergoing daily triaging including temperature check, working in teams, disinfection routine and adherence to a clinic-specific code of conduct (Supplementary data) which later incorporated ESHRE and Indian advisory [9,11]. Triage questionnaire went through periodic review and changes to meet the demands of the evolving pandemic [supplementary data; 9,16,17]. Two teams, each consisting of at least one IVF nurse, a clinician, an anesthetist and two embryologists skilled in performing ICSI, vitrification and warming were created. Members of both teams resided in different geographic areas to minimize the chance of members from both teams being in the same containment zone and getting quarantined simultaneously [18]. Time required to comply with all the steps and for procurement of appropriate standard personal protective equipment (PPE) was tracked on a daily basis.

### Patient recruitment

Once the clinic preparedness was ensured, couples wishing to commence their treatment were contacted. Through tele-consultation, treatment plan, code of conduct, available data on pregnancy outcomes in COVID-19 and need for the couple to self-isolate from two weeks prior to initiation of ovarian stimulation through the duration of treatment were discussed. The reasons for consenting or not agreeing to IVF procedures were recorded. Consent forms were sent electronically to the couples and a follow-up tele-consultation done to clarify any doubts. This was followed by an ‘in-person’ consultation to reinforce the above information, ensure couple’s understanding of code of conduct, possibility of cancellation if the pandemic worsened locally or if either or both contracted or suspected of having COVID-19, alternative arrangement if any personnel from clinic got infected, financial implications, and to sign consent forms. Importantly, they were counseled regarding the current lack accessibility to COVID testing for asymptomatic people in the local area. Semen cryopreservation if not previously done, was performed during their visit to hospital and post-wash samples were stored in a dedicated cryocan.

Amongst those who wished to start IVF, priority was given to couples with wife’s age >35 years, proven or expected poor ovarian reserve [anti-Mullerian Hormone (AMH) <1.2ng/ml and antral follicle count (AFC) <8], and fertility preservation (malignant and benign conditions).

Those with apprehension about treatment during pandemic, financial concerns or without safe transport were advised to defer treatment. Some were referred to nearby clinics to avoid undue delay due to travel restrictions.

### Treatment

Clinic 1 utilized both agonist and antagonist protocols, IVF or ICSI, based on ovarian reserve markers and semen parameters respectively; and selective embryo freezing. Clinic 2 utilized antagonist protocol, ICSI and elective freezing of all embryos in fresh cycles. Starting dose of gonadotropin depended on the ovarian reserve markers in both the clinics. All women received human Chorionic Gonadotropin (HCG) as trigger. Frozen embryo transfer (FET) was performed in hormone replacement therapy (HRT) cycles following mid-luteal pituitary downregulation in both clinics. Anaesthesia for OPU depended on the routine practice in each clinic. Women in Clinic 1 were counseled and offered CSA with intravenous (IV) Midazolam and Fentanyl. Propofol was available for use if pain control was not satisfactory. Pain score was documented on a graphic rating scale (GRS) of 10 cm length extending from no pain through mild, moderate to severe pain [19]. In Clinic 2, GA with IV Propofol and Fentanyl along with intubation box was used during OPU. Duration of OPU was documented. Sequential embryo assessment was performed in Clinic 1 while uninterrupted single step culture was utilized in Clinic 2. Those undergoing embryo transfer received standard luteal support with vaginal progesterone.

### Key performance Indicators (KPI)

Reference Indicators (RIs), Performance Indicators (PIs) and Key Performance Indicators (KPIs) to assess the teams’ performance during the pandemic were done according to ESHRE Vienna consensus criteria [20]. 18 out of the total 19 recommended parameters relevant to both the clinics involving ovarian stimulation, fertilization and post-fertilization laboratory parameters were documented.

### Statistical analysis

As this study reports initial experience after resuming IVF work during COVID-19 pandemic, a sample size calculation was not performed. Data for both the clinics is pooled and represented. The data was prospectively maintained in Microsoft excel. Where indicated, the inter-clinic parameters were compared using Student’s t-test and the results expressed as mean ± standard deviation (SD). Cumulative data is expressed as actual numbers and percentages.

## Results

### Time taken for prepare the clinic and staff according to new norms

Time taken to achieve safe functional preparedness is shown in Table 1. Amongst the laboratory requirements, availability of fresh stock of media and consumables took the longest, followed by fresh stock of gonadotropin injections. The operation theater preparedness was uninterrupted. Most of the COVID-19 specific requirements could be organized within a week’s time, while achieving appropriate social distancing measures within the clinics took more than 2 weeks. One nursing staff declined working for the fear of COVID-19 while majority immediately showed their willingness to work and resumed duties upon availability of safe transport organized by the clinics. The timeline for preparedness was similar in both clinics.

**Table 1:** Time taken for resumption of services in the IVF clinics post lockdown imposed due COVID-19 pandemic.

| Services | Time taken for re-initiation since lockdown |
| --- | --- |
| **Laboratory preparedness** |  |
| LN2 supply | Uninterrupted |
| Calibrated CO<sub>2</sub> cylinders | Already available |
| Daily logs | Uninterrupted |
| Availability of fresh culture media | 45 days |
| Availability of IVF laboratory consumables | 45 days |
| Availability of Andrology laboratory consumables | 2 days |
| Establishing contact with equipment suppliers | 8 days |
| Availability of laboratory disinfectant | Continuous |
| **Operation theater preparedness** |  |
| Availability of fresh stock of anesthetic medications | Uninterrupted |
| Availability of anesthetic gases | Uninterrupted |
| Consumables for routine procedures | Uninterrupted |
| **Preparedness for Superovulation** |  |
| Gonadotropins, agonists and antagonists for superovulation | 30 days |
| Oral / transdermal estrogens; progesterone preparations | 7 days |
| **COVID-19 specific preparedness** |  |
| Procuring N95 masks and other PPE | 7 days |
| Organizing the clinics for triage, isolation area, & patient movement | 2 days |
| Developing tele-consultation questionnaire | 1 day |
| Establishing tele-consultation system | 5 days |
| Sanitization of hospital every 4 hours | Immediate |
| **Staff preparation for COVID-19 specific requirements** |  |
| Number of staff willing to work | 31/32 (92.3%) |
| Number of staff eligible to work after completing triaging | 31 |
| Number of staff compliance with daily triage (since 25/3/20) | 100% |
| Safe transport services for staff | 2 days |
| Staff compliance for wearing mask | Immediate |
| Compliance with hand sanitization | 2 days |
| Compliance with appropriate PPE | 7 days |
| Training for social distancing practices | 17 days |
| Training for obtaining patient consent for tele-consultation | 2 days |

CO<sub>2</sub> = Carbon dioxide; LN2 = Liquid nitrogen; PPE = Personal Protective Equipment.

### Operationalization and outcomes of telemedicine services

In absence of SARS-CoV-2 testing services for asymptomatic individuals, the triage questionnaire played an important role and needed regular modification due to evolving situation. Triage questionnaire and telemedicine questionnaire were prepared within a week and the services could be implemented immediately in both the clinics (Table 1 and supplementary data). 169 couples were consulted by telemedicine based on the pre-lockdown appointment logs (Table 2). Only 30% of patients wished to avail services for ART post resumption services. Lack of easy access to clinics was the most prominent reason to delay treatment, followed by financial constraints (Table 2).

**Table 2:** Outcomes of tele-consultation by the IVF clinics after resumption of services post lockdown imposed due to COVID-19 pandemic.

| **Patient details** | **No. of couples** |
| --- | --- |
| Total number of couples administered with tele-consultation questionnaire | 169 |
| Number of couples agreeing to / eligible for treatment at tele-consultation | 52 (30.8%) |
| Number of patients declining/ advised against treatment at tele-consultation | 117 (69.2%) |
| Reasons for declining: |  |
| Fear of pandemic | 11 (9.4%) |
| Financial reasons | 24 (20.5%) |
| Lack of access to clinic | 72 (61.5%) |
| Presence of co-morbidities | 09 (7.6%) |
| Number of patients transferred to other clinics for accessibility | 01 (0.85%) |

### Characteristics of couples who underwent ART

Fifty-two couples underwent treatment upon resumption of ART services. All couples complied with new norms, triage during every visit, underwent isolation (self-reported) for two weeks prior to and during treatment and agreed to freeze all embryos (and cancel embryo transfer) if advised due to any change in pandemic scenario. Table 3 shows that majority of the couples-initiated treatment in view of medical urgency (fertility preservation, expected or proven POR). However, a proportion of infertile couples chose to go through treatment without delay due self-perceived urgency or preparedness.

**Table 3:** Indications for assisted reproduction in infertile couples who underwent treatment after resumption of services post lockdown imposed due to COVID-19 pandemic.

| **Indication for IVF** | **No. of couples**<br>**(n=52)** |
| --- | --- |
| Fresh IVF cycles | 28 |
| Frozen embryo transfer cycles | 24 |
| Fertility Preservation | 2 (3.8%) |
| Age >35 years | 15 (28.8%) |
| Poor ovarian reserve | 12 (23.1%) |
| Severe male factor | 15 (28.8%) |
| PCOS achieving desired weight reduction | 1 (1.9%) |
| Couples' choice (due to career) | 7 (13.5%) |

PCOS = Polycystic Ovarian Syndrome

### IVF Treatment Details

The mean age of women undergoing ART was 32.3±3.5 years. There was no incidence of ovarian hyperstimulation syndrome (OHSS) or COVID-19 and one cycle was cancelled due to no response. Different anaesthesia techniques used for OPU show a significantly less time for OPU with CSA compared to GA for retrieval of similar number of oocytes (Table 4). Further, mapping of pain score on a GRS revealed high acceptance rate of CSA (Table 4). The overall clinical pregnancy rate per cycle was 41.7% in fresh cycles and 48.1 % in frozen cycles

**Table 4:** Anaesthesia and oocyte pick up (OPU) details from both clinics after resumption of services post lockdown imposed due to COVID-19 pandemic.

| Parameter | Protocol 1 | Protocol 2 | P value |
| --- | --- | --- | --- |
| Anaesthesia used for OPU | CSA | GA |  |
| Number of patients | 14 | 13 |  |
| Duration of OPU (minutes) | 17.0±3.1 | 23.7±10.9 | 0.03* |
| Pain score during OPU as documented on a GRS | None: 1<br>Mild: 12<br>Moderate: 1 | - | - |
| Number of oocytes (mean ± SD) | 12.6±7.4 | 14.5±7.5 | 0.5 |

CSA = Conscious Sedation and Analgesia; GA = General Anaesthesia; GRS = Graphic Rating Scale; OPU = oocyte pick up
\*P = significant; value is by student's t test.

### KPIs after resumption of services

Table 5 shows RIs, PIs and KPIs of the clinics during the pandemic, in comparison to historic data of 3 months prior to closure of the clinic (October 2019 to December 2019). As evident, these indicators of clinics’ performance matched the historic data and were above the competency value or approached benchmark values as per ESHRE Vienna Consensus.

**Table 5:** Reference / Performance / Key performance indicators in IVF clinics after resumption of services post lockdown imposed due to COVID-19 pandemic.

| Reference/Performance/<br>Key Performance<br>Indicators | Values during<br>study period | Historic data | Competency value<br>(Vienna<br>Consensus) | Benchmark<br>Value<br>(Vienna Consensus) |
| --- | --- | --- | --- | --- |
| Time period | Apr 2020 – July 2020 | Oct 2019 – Dec 2019 |  |  |
| Number of patients | 52 | 117 |  |  |
| **Reference Indicators** |  |  |  |  |
| Proportion of oocytes recovered | 366/389 (94.1%) | 822/875 (93.9%) | - | 80-95% of follicles |
| Proportion of MII oocytes | 265/350 (75.7%) | 503/670 (75.1%) | - | 75-90% |
| **Performance Indicators** |  |  |  |  |
| Post-preparation sperm motility | 90% | 90% | 90% | ≥95% |
| Polyspermy in IVF | 0 /15 (0%) | 5/152 (3.3%) | - | <6% |
| 1 PN in IVF | 0 /15 (0%) | 6/152 (3.9%) | - | <5% |
| 1 PN in ICSI | 3/265 (1.1%) | 10/503 (1.9%) | - | <3% |
| Good blastocysts | 40/82 (48.8%) | 48/107 (44.8%) | - | ≥40% |
| **Key Performance Indicators** |  |  |  |  |
| ICSI damage rate | 25/285 (8.8%) | 59/503 (11.7%) | <=10 | <=5 |
| ICSI –normal fertilization | 187 /285 (65.6%) | 310/503 (61.6%) | ≥65 | ≥80 |
| IVF –normal fertilization | 10/15 (66.7%) | 98/152 (64.5%) | ≥60 | ≥75 |
| Failed fertilization in IVF | 0 | 0 |  | <5 |
| Cleavage rate | 87/93 (93.5%) | 328/408 (80.3%) | >=95 | >=99 |
| Day 2 embryo development rate | 63/93 (67.7%) | 228/408 (55.3%) | >=50 | >=80 |
| Day 3 embryo development rate | 134/197 (68%) | 168/408 (41.1%) | >=40 | >=60 |
| Blastocyst development rate | 73/156 (46.7%) | 52/107 (48.5%) | >=40 | >=60 |
| Implantation rate (Day 3) | 2/6 (33.3%)* | 18/70 (25.7%) | >=25 | >=35 |
| Implantation rate (Day 5) | 4/6 (66.6%)* | 12/25 (48%) | >=35 | >=60 |
| Blastocyst cryosurvival rate | 25/26 (96.2%) | 50/52 (96.1%) | >=90 | ./=99 |
| Implantation rate of vitrified and warmed embryos (Day 5) | 12/25 (48%) | 26/50 (52%) | - | - |

Values are expressed as numbers (%).
\* Data is from embryo transfer in fresh IVF / ICSI cycles
ICSI = Intra-cytoplasmic sperm injection; IVF = in-vitro fertilization; MII = Meiosis II, PN = Pronucleus

## Discussion

To our knowledge this is the first study to evaluate the three important factors that influence successful resumption of IVF during the COVID-19 pandemic – clinic preparedness, informed decision of patients to go through treatment, and KPIs of the clinics. The study highlights the need for a multipronged approach cognizant of local, national and international scenario for successful resumption of ART services under the newly defined norms and code of conduct to mitigate the risk of SARS-CoV-2 infection in a hotspot region of India.

Resumption of ART services during pandemic is both a clinical and social dilemma [3,12]. Infertility continues to be the top stressor even in the midst of the pandemic and delaying treatment may add further stress to such couples [21]. Whether psychological stress affects IVF outcome remains a controversial issue [22,23]. However, as the pandemic continues to disrupt routine life, it may become important for those who have not been able to access fertility services [12,24]. While short-term delay may have no negative impact on IVF outcomes, this has to be balanced against impact of prolonged delays on population dynamics and age related decline in live births [25-27].

One of the first key challenges we faced was the interrupted supply of perishables such as IVF culture media and injectables for ovarian stimulation. While other supplies and functionalities were uninterrupted, procurement of perishables took the longest. This was understandably the consequence of restricted import and regional transport services due to lockdown. Our results indicate the need for an anticipatory planning, close monitoring of the ordering and purchase routines and co-ordination not only at the clinic level but with distributors, and manufacturers as well, both for completion of on-going cycles and for smooth restarting [28]. Another major requirement for service resumption in an IVF clinic is the staff adapting to the new norms [28]. Contrary to common expectation, most laboratory, paramedical and support staff were willing to undergo specific training and resume work even in the midst of the pandemic. This is heartening for both clinicians and patients. Once safe transport was organized, they promptly concentrated on training; and adaptation to new practices was achieved within a week with the exception of social distancing measures. The staff maintaining at least 2 meter distance from each other while working and minimizing ‘in-person’ interactions were hard to achieve and took more than two weeks to ensure compliance.

To minimize ‘in-person’ visits to the clinic and provide appropriate clinical services, use of telemedicine has been widely advised [2,3,5,6,28]. We devised a tele-questionnaire that spanned questions not just about the medical problems but also provided information on clinic preparedness and obtain an understanding telephonically if the couples would qualify triaging. Of those who deferred treatment, nearly 62% did so due to lack of appropriate transport to access the clinic. While nearly 20% declined stating financial reasons, only 9% of couples deferred due to fear of pandemic. The findings highlight the possibility of financial crises the society is facing affecting IVF services eventually [29]. In addition to medically urgent IVF, informed decision by patients to initiate treatment also amounts to a valid indication. Interestingly nearly 42% underwent treatment because they identified lockdown as an opportunity to improve their lifestyle which otherwise was a challenge for them due to heavy work schedule. It will be important for reproductive medicine specialists to recognize this demand, which by no stretch of imagination is a medical emergency but a socially justified emergency [12,29-32].

Triaging of couples and team members and adherence to code of conduct as an integral strategy does instill a level of discipline at the clinics [9,11,15,16]. This is a crucial step towards judicious utilization of resources while striving to provide optimal services. Scientific societies differ in their recommendations of relative roles of triaging and testing for SARS-COV-2 [16,33]. Further, as India had closed its international borders since March 2020, the questionnaire pertaining to international travel became irrelevant. Our experience highlights that triaging is an evolving concept based on local needs and situations, to ensure patient and staff safety. Even though we could successfully resume IVF, an upward trajectory of COVID-19 demands continued vigilance. While the testing services are still largely prioritized to symptomatic patients and those with high-risk exposures, availability and accessibility for testing are increasing in India [34]. As the time span of pandemic increases, understanding of utility, constraints and limitations of different tests for SARS-CoV-2 is increasing [35,36]. Failure to appreciate the lacunae of various tests and undue reliance on them may prove to be detrimental for the ART program [33,35].

OPU is the only step in IVF during which considerable time is spent in close proximity to patients. Many different types of anesthesia or analgesia are equally effective in achieving patient comfort during OPU [37,38]. The findings of this study show that the procedure time is significantly less with CSA compared to GA for retrieval of similar number of oocytes. Reduced procedural time and no intervention for airway maintenance have important implications in this pandemic with a respiratory virus. Considering that the pain score was low and patient satisfaction was high, this is a useful strategy to mitigate the infection risk to health care professionals, in addition to use of appropriate PPE.

Success of ART depends on the clinical and laboratory performance indicators, which benchmark the clinic’s performance. The additional challenges for IVF during this pandemic are the anxieties, added responsibilities and concerns faced by the IVF team adversely affecting the KPIs [20,28]. We evaluated these less often reported parameters and found that despite new and diverse responsibilities the stimulation and laboratory parameters were consistently above the competency level or approaching benchmark level. In addition, the KPIs were comparable to the pre-pandemic values and provide an objective evidence of clinical and laboratory performance during the pandemic. This is reassuring and will encourage other clinics to resume services for the benefit of the patients.

While the ART fraternity debates on resumption of services and struggles with strategies, one must keep in mind that ART pregnancies may not be absolutely protected from SARS-CoV-2 [39-41]. There is evidence of the presence of SARS-CoV-2 receptors in both female and male genital tract and gametes; the virus is detected in semen and may cause gonadal dysfunction [42-45]. Although, it is believed that zona pellucida acts as a barrier to prevent infection of embryo, the cells of the developing human blastocysts, trophectoderm and early placenta are permissible sites of SARS-CoV-2 infection [46-49]. A large proportion of carriers of the virus are asymptomatic, there is a high human-to-human transmission rate and the virus can survive on surfaces for unusually long periods [50]. Clinicians, laboratory and paramedical staff must be constantly aware of this and act appropriately. Post conception, the pregnancies in COVID-19 is another controversial area. Albeit small in number, there is definitive evidence of placental infection and vertical transmission of SARS-CoV-2 [51-55]. While there is no evidence to suggest that the ART pregnancies are additionally susceptible or protected from the ill effects of COVID-19, the possibility of vertical transmission from an IVF clinic may be negligible [57]. It is important that the couples are informed of the current evidence of COVID-19 on pregnancy prior to opting for ART.

In summary, this is the first study describing an experience of reopening IVF services during this pandemic. It shows the diverse areas to be addressed while achieving functionality of the clinics. We show that 1) the preparedness of labs and hospital setup may not be time consuming but the supplies need to be ensured, 2) there will be need for an individualized approach for selecting couples to undergo IVF 3) the performance of clinicians and embryologists in the face of uncertainties and anxieties due to the pandemic may not be compromised if adequate measures are taken and training provided. To this end, whether this experience will matter to all the clinics globally is debatable but we are certain that the challenges faced by us will be applicable to most clinics in low to middle income countries. Through this communication we wish to indicate that as the role of SARS-COV-2 testing in IVF remains unclear and while access to testing is restricted, it is the important to develop clinic specific triaging norms to resume services and if implemented diligently it is possible to achieve IVF pregnancies even during the pandemic.

## Data Availability

The data for this manuscript is available in the central repository of Shreyas Hospital under IVF resumption during COVID and the authors will share the data whenever necessary.

## Acknowledgements

The authors gratefully acknowledge the assistance provided by Miss Rushita Vaghasia and Miss Pranita Bawaskar, Nagpur test tube baby centre, Nagpur, for data compilation.

